# Prospective comparison of saliva and nasopharyngeal swab sampling for mass screening for COVID-19

**DOI:** 10.1101/2020.09.23.20150961

**Authors:** Mathieu Nacher, Mayka Mergeay-Fabre, Denis Blanchet, Orelie Benois, Tristan Pozl, Pauline Mesphoule, Vincent Sainte-Rose, Véronique Vialette, Bruno Toulet, Aurélie Moua, MONA Saout, Stéphane Simon, Manon Guidarelli, Muriel Galindo, Barbara Biche, William Faurous, Fanny Abad, Aniza Fahrasmane, Devi Rochemont, Nicolas Vignier, Astrid Vabret, Magalie Demar

## Abstract

Current testing for COVID-19 relies on quantitative reverse-transcriptase polymerase chain reaction from a nasopharyngeal swab specimen. Saliva samples have advantages regarding ease and painlessness of collection, which does not require trained staff and may allow self-sampling. We enrolled 776 persons at various field-testing sites and collected nasopharyngeal and pooled saliva samples. 162 had a positive COVID-19 RT-PCR, 61% were mildly symptomatic and 39% asymptomatic. The sensitivity of RT-PCR on saliva samples versus nasopharygeal swabs varied depending on the patient groups considered or on Ct thresholds. There were 10 (6.2%) patients with a positive saliva sample and a negative nasopharyngeal swab, all of whom had Ct values<25. For symptomatic patients for whom the interval between symptoms onset and sampling was <10 days sensitivity was 77% but when excluding persons with isolated Ngen positivity (54/162), sensitivity was 90%. In asymptomatic patients, the sensitivity was only 24%. When we looked at patients with Cts <30, sensitivity was 83% or 88.9% when considering 2 genes. The relatively good performance for patients with low Cts suggests that Saliva testing could be a useful and acceptable tool to identify infectious persons in mass screening contexts, a strategically important task for contact tracing and isolation in the community.

## Introduction

Current testing for COVID-19 relies on quantitative reverse-transcriptase polymerase chain reaction (RT-qPCR) from a nasopharyngeal swab specimen.(1) Nasopharyngeal sampling requires human resources and training, personal protective equipment and swabs, and time, generating testing bottlenecks and potential exposure to transmission at crowded testing sites. Moreover, the unpleasantness of the procedure and the long waiting delays may dissuade some persons to get tested or to repeat tests when they are negative. There is an urgent need for innovative testing strategies to rapidly identify cases, reduce waiting delays, and facilitate mass screening. Saliva samples have advantages regarding ease and painlessness of collection, which does not require trained staff and may allow self-sampling. The comparison of real time PCR results on salivary and nasopharyngeal samples has shown discrepancies between studies, with most finding greater sensitivity and lower RT-PCR Cts in nasopharyngeal swab samples(2-4) whereas others found greater sensitivity in saliva samples(5, 6). The sources of variation may have been the study population (hospitalized patients versus screening of contacts or mildly symptomatic patients), saliva collection techniques and timing, conditioning and delays in processing raw saliva samples, or differences in the RT-PCR techniques used.

We here report the first prospective study of the performance of saliva testing compared to nasopharyngeal swabs in a field context of mass screening in French Guiana.

## Methods

### Context

This French territory neighboring Amapa state in Brazil has been highly impacted by COVID-19 with 3.2% of the population having had a confirmed infection, notably among the poorest populations.(7) In this context, testing and tracking were implemented throughout the epidemic, testing tents and mobile testing teams including the remote health centers, the Red Cross, Médecins du Monde, and the reinforcements from the Réserve Sanitaire were coordinated by the regional health agency to investigate around clusters of cases. The testing efforts for this small population peaked to nearly 0.5% of the population screened in a day.

### Study conduct

Between July 27^th^ and September 10^th^, we prospectively enrolled consecutive, persons aged 3 years or more with mild symptoms suggestive of COVID-19 and high-risk asymptomatic persons at various testing sites and mobile testing brigades in French Guiana reaching remote sites up to 240 km in the Amazonian Forest. During screening missions, mobile teams, consisting of Healthcare personnel (doctors, nurses) were coordinated by the Health Regional Agency of French Guiana, targeting villages, neighborhoods, where the virus was circulating collected persons often out of doors or in health centers. These mobile teams were made up of staff from the Red Cross, Médecins du Monde, the Cayenne hospital PASS and the health reserve. Team travel was coordinated and decided by the health regional agency of French Guiana each week during a weekly update and was guided the knowledge of clusters or a screening campaign in different neighborhoods. Inclusion criteria were: males or females with an indication to perform a COVID diagnostic test (symptomatology, contact case, systematic screening, etc.), aged at least 3 years old. Non-inclusion criteria were refusal of the patient or his legal representative, person taking treatments that reduce salivary volume (anticholinergic activity), impossibility of carrying out the NPS, and persons under guardianship or curatorship, or placed under protective measures. All study participants were enrolled and sampled in accordance with the protocol. An investigator explained the objectives of the study and obtain the consent of the patient or his legal representative. The form was completed by the investigator or a person delegated by the investigator. The trained nurse present during the testing mission performed the nasopharyngeal swab collected the salivary sputum sample in a urine container. A trained agent carried out a short questionnaire. At the end of each day, all completed forms and samples were sent to Cayenne hospital and stored at 4°C before analysis. Samples and participant information were non-individually identifiable and collected with a unique identifying number.

### Laboratory analysis

The same technique was used for the 2 samples throughout the study: the QIAsymphony and GeneFinder kit, a Real-time PCR assay. GeneFinder™ COVID-19 detects SARS-CoV-2 by amplification of RdRp gene, E gene and N gene according to WHO’s recommended protocol. Viral nucleic acid was extracted by using the QIAamp DSP viral kit on the QIAsymphony RGQ, an integrated fully automated nucleic acid extraction (chemical lysis and paramagnetic bead binding) and sample preparation platform (Qiagen GmbH, Germany). The real-time PCR assays for SARS Cov2 were performed with an Applied 7500 cycler (Thermofisher) with the Genefinder kit (Ellitech group) that could detect the Ngen, RdRP and E gen. As the Nucleic acid extraction methods could affect the results of viral nucleic acid amplification tests, we treated the couple saliva-nasopharyngeal specimens with the same method and most of the time in the same series, the eluates were obtained from 200μl of specimens (300 μL minus 100 μL dead volume). The remainder of each sample was divided into paired aliquots kept in a biorepository for further studies evaluating new screening tools.

### Statistical analysis

Statistical analysis was performed using STATA® 16 (Stata corporation, College Station, Texas, USA). Cross tabulations considering different subgroups was performed. We considered the specific genes for SARS-Cov2 RdRp and Ngene to calculate different Ct categories.

### Ethical

The protocol received ethical approval from the Comité de Protection des Personnes under the number 2020-A02009-30/SI:20.07.07.54744.

## Results

We included 776 patients, 162 (20.9%) of whom had a positive result (152 nasopharyngeal and 86 saliva) (Figure 1). The sex ratio (M/F) was 1.6, the mean age was 40 (standard deviation=16.8). Overall, 61% were mildly symptomatic and 39% were asymptomatic.

**Fig 1a.**
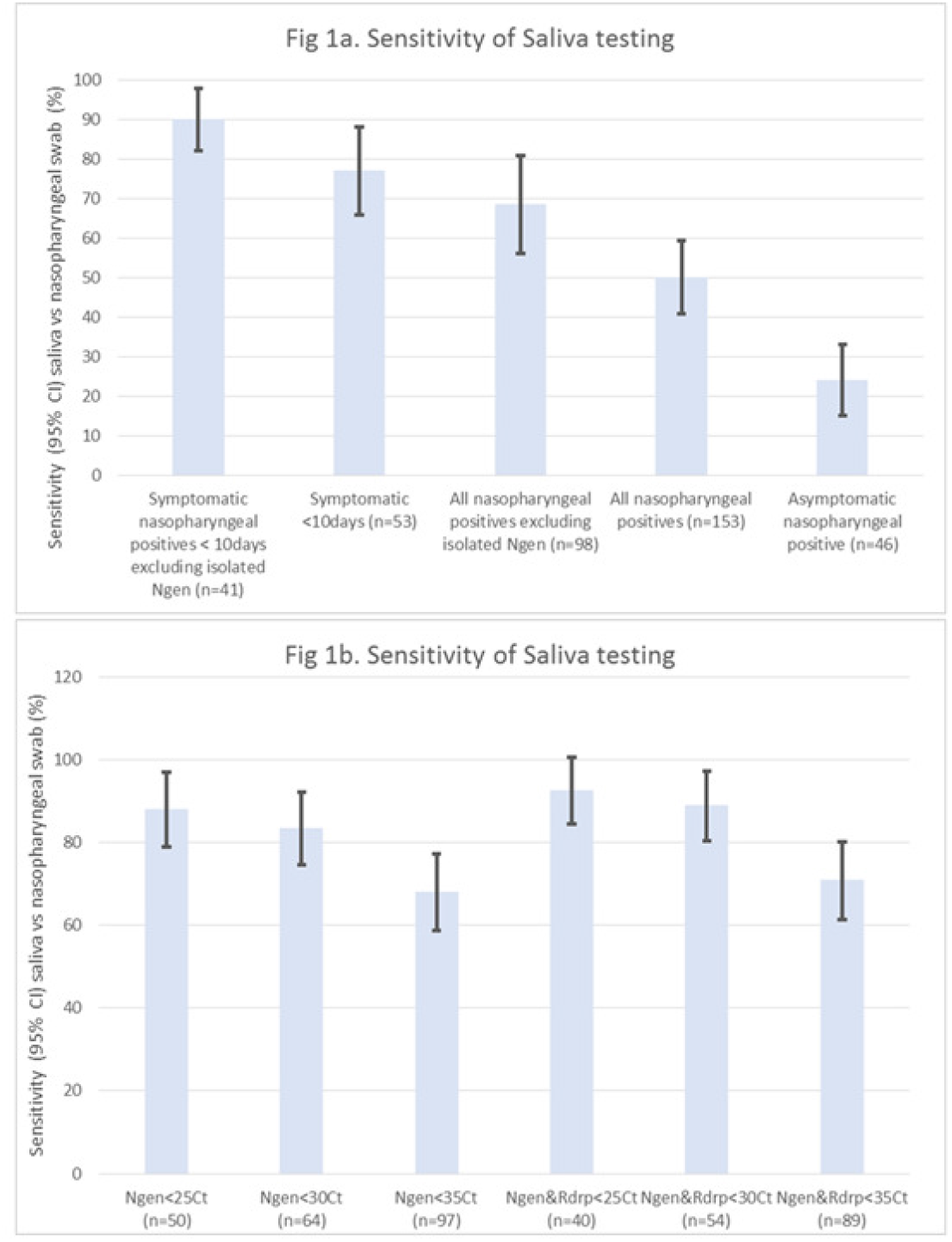
Sensitivity of saliva testing versus nasopharyngeal swabs for RT-PCR for different groups in a community screening context; Fig 1b. Sensitivity of saliva testing versus nasopharyngeal swabs for different RT-PCR Cts in a community screening context.

For symptomatic patients, 84% had a symptoms onset <10 days, and 4% were hospitalized within 2 weeks after inclusion. The sensitivity of RT-PCR on saliva samples versus nasopharygeal swabs varied depending on the patient groups considered (Fig 2a) or on Ct thresholds (Fig 2b). There were 10 (6.2%) patients with a positive saliva sample and a negative nasopharyngeal swab, all of whom had Ct values<25. For symptomatic patients for whom the interval between symptoms onset and sampling was <10 days sensitivity was 77% but when excluding persons with isolated Ngen positivity (54/162), sensitivity was 90%. In asymptomatic patients, the sensitivity was only 24% (Fig 2a). Recent studies have argued that transmission potential -estimated by the capacity to infect cell cultures-was restrained to those with low Cts (8, 9), a proxy for high viral load. When we looked at patients with Cts <30, sensitivity was 83% or 88.9% when considering 2 genes. Figure 3 shows a trend for fanning towards the higher Ct values the nasopharyngeal versus saliva sample scatterplots for the different genes amplified by RT PCR. The positive predictive value of saliva samples was 88.4% and the negative predictive value was 88.9%.

**Figure 2.**
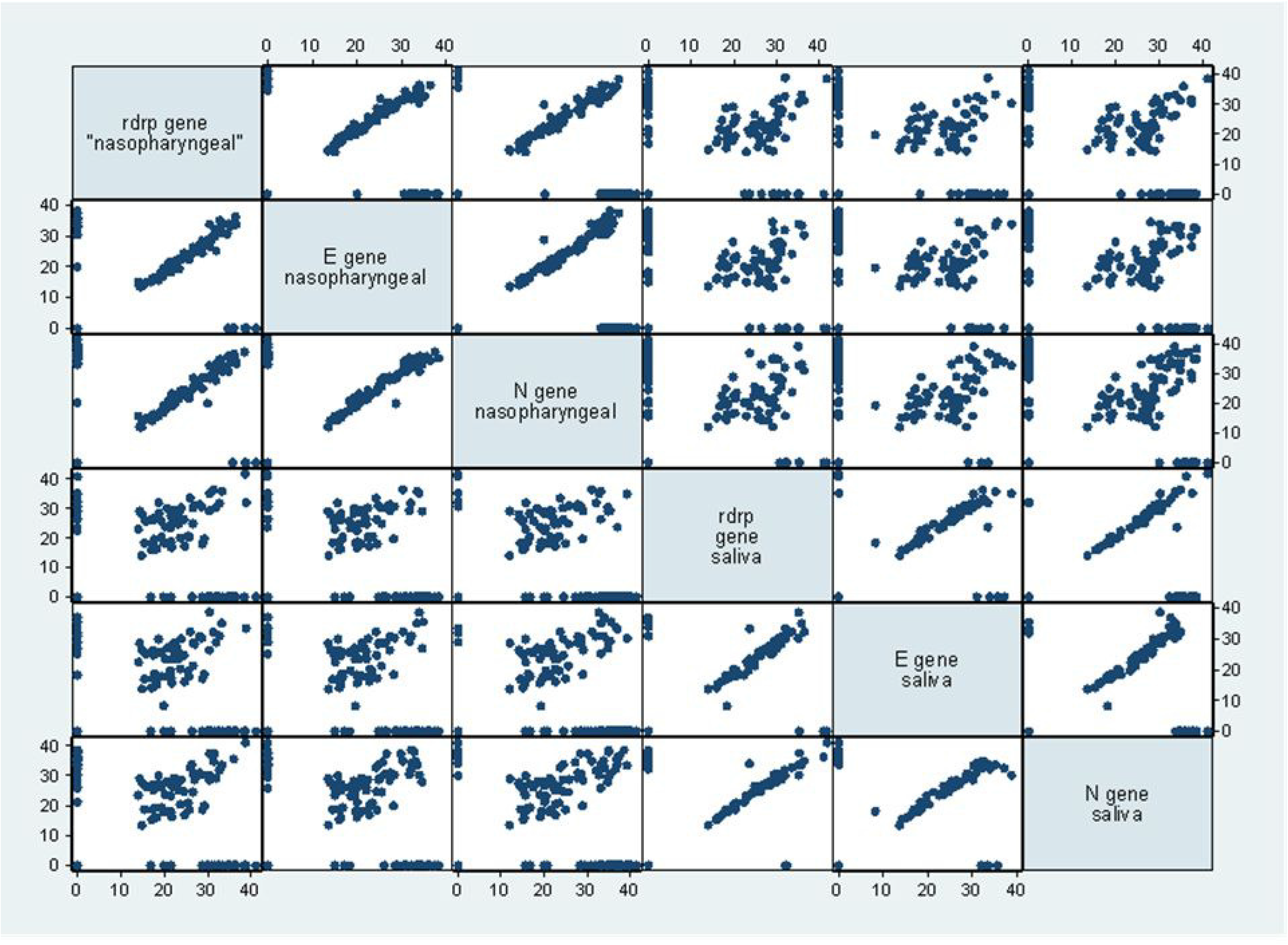
Scatterplot matrix for the Ct of different genes in the nasopharyngeal and saliva samples. There was a “fanning” pattern with greater dispersion at higher Ct values for different genes in the nasopharyngeal and saliva samples.

## Discussion

Contrarily to 2 studies suggesting a greater positivity rate for saliva(5, 6), we observed that saliva testing was less sensitive than nasopharyngeal swabs. Whereas most studies were hospital-based collecting saliva in the early morning before mouth rinsing and breakfast, our study was a screening study that was performed in difficult field conditions targeting hard to reach populations after breakfast and teeth brushing, moreover out of doors in a tropical context. The poor sensitivity on asymptomatic positive nasopharyngeal swabs was presumably also linked to the inclusion of non-infectious patients in the denominator. The relatively good performance for patients with low Cts suggests that Saliva testing could be a useful and acceptable tool to identify infectious persons in mass screening contexts, a strategically important task for contact tracing and isolation in the community. With the considerable testing bottlenecks, although the time and workload for RT-PCR itself would be similar, alleviating the workload and shortening the sample collection time would be improvements that could reduce waiting times to get tested and human-resource costs. The sensitivity saliva samples for asymptomatic persons seemed insufficient but without any temporal indication about the onset of infection, it should be further studied by Ct values with a larger sample size. In view of the present results the French Health authorities have officially declared that saliva testing may be used on symptomatic patients only.(10)

## Data Availability

data can be made available after further authorization from the commission nationale informatique et libertes as required by french law

## Acknowledgements

We wish to express our gratitude to the directions and personnel of the Agence Régionale de Santé de Guyane, Centre Hospitalier de Cayenne, the Croix Rouge Française, Médecins du Monde, the Permanence d’Accès aux Soins de Santé, the Réserve Sanitaire, Santé Publique France, the Centres Délocalisés de Prévention et de Soins and numerous Health mediators, and REACTing

---

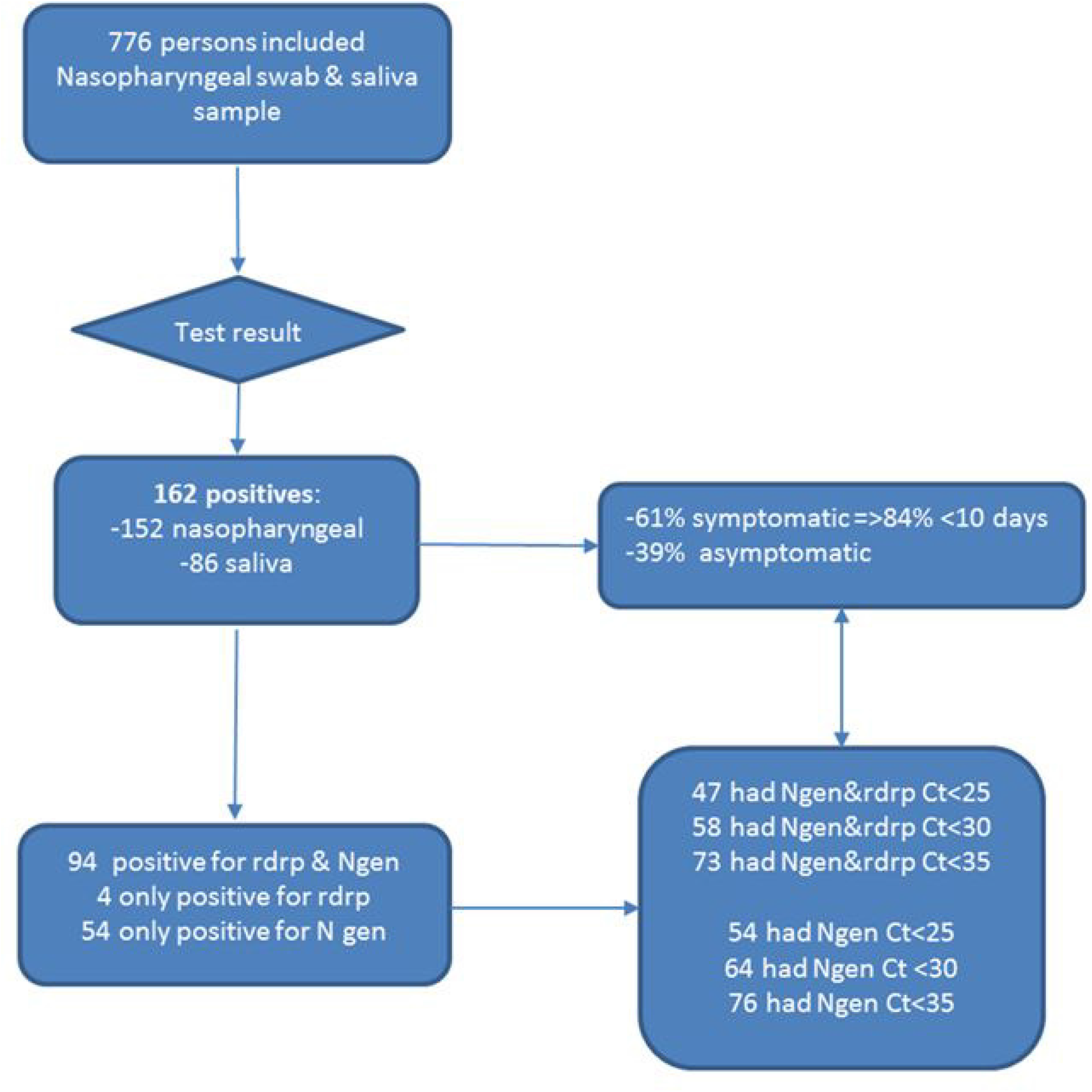

